# Establishment of an External Quality Assessment Scheme for Clinical Parasitology Diagnostics in India: Four years of experience (2022-2025)

**DOI:** 10.64898/2026.07.22.26358636

**Authors:** Renita Ruth, Selvi Laxmanan, Ramya Madhavan, Tintu Varghese, Peter L Chiodini, Jaap J. van Hellmond, Sudhir Babji, Sitara Swarna Rao Ajjampur

## Abstract

**Introduction:** Microscopy remains an important ‘catch all’ diagnostic tool for parasite detection in clinical settings. Although relatively low-cost in endemic settings, it requires sustained technical skills. To enhance microscopy-based diagnostic capabilities in India, a parasitology External Quality Assessment Scheme (EQAS) was launched in 2022.

**Materials/Methods:** An online survey was conducted to plan the diagnostics scope for a clinical parasitology EQAS in India. In the EQAS, well-characterized proficiency testing material was distributed biannually, with two challenges/cycle and scoring provided based on established criteria. After three years, an online feedback survey was conducted to evaluate the perceived effectiveness of the scheme.

**Results:** The diagnostic scope assessment (n=78 laboratories) indicated microscopy as the commonest technique for parasite identification (100%) and molecular diagnostics are sparsely available (3.8%). Of the eight PT cycles conducted, high competence (>80%) was observed for endemic parasites, *Giardia duodenalis* (90% in 2022 & 97% in 2025), *Echinococcus granulosus* (96%), *Trichuris trichiura* (95%), *Taenia spp.,* (88.6%), *Strongyloides stercoralis* (87%), *Hymenolepis nana* (84%), *Ascaris lumbricoides* (82.6%), *Cystoisospora belli* (81%), and *Cryptosporidium spp.* (80%). Challenges noted were in the identification of uncommon or non-endemic parasites - *Paracapillaria philippinensis* (58%), *Clonorchis sinensis* (72%), *Hymenolepis diminuta* (63%) and in samples with two parasites, only 27% reported both accurately. Feedback survey indicated >95% satisfaction with reporting and educational materials among respondents, and 90% requesting for 3 PT cycles/year.

**Conclusion:** The parasitology EQAS program tailored for Indian laboratories led to measurable improvement in diagnostic performance, and participants’ feedback indicated a positive impact on parasitology diagnostics and education.

## INTRODUCTION

Advances in laboratory techniques have transformed parasitology diagnostics, expanding access to accurate and timely testing, improved detection rates both in the clinical setting and in public health programs, and enabled identification and characterization of emerging parasites and drug resistance [1–2]. Despite these technological advancements, traditional microscopy and morphology- based analyses remain the cornerstone for parasite identification across various diagnostic settings [3]. In resource-limited settings and at the point-of-care, broad applicability as a ‘catch-all’ diagnostic, rapid turnaround time, and minimal equipment requirements make microscopy the most accessible and cost-effective diagnostic tool. Preference for non-microscopy methods such as rapid immunochromatographic tests (rapid diagnostic tests, RDT) intended for public health programs can lead to a decline in technical skills among clinical parasitologists and laboratory technicians. Sustaining these competencies is necessary to provide diagnostic quality and preserve these critical skills among future clinical parasitologists, especially in highly endemic and low resource settings [3–4]. This also applies to future AI based digital pathology tools for parasite identification where a ‘human-in-the-loop’ in the clinical setting would be critical [5]. One of the most effective ways to support and improve performance in clinical laboratories is by implementing an External Quality Assessment Scheme (EQAS). Studies conducted across different settings have demonstrated that EQAS participation can significantly enhance microscopy skills, thereby improving individual diagnostic performance, and lead to accreditation and overall improved laboratory performance and practice [6–9].

In India, there is limited information available on clinical parasitology diagnostics – on utilisation, availability, methods and practices in the hospital setting [10]. There is recognition of the need for an affordable and accessible structured parasitology EQAS program with the diagnostic scope tailored to available clinical parasitological diagnostics in India. To address this gap, a national-level online survey was conducted to assess clinical parasitology diagnostic practices and methods available, following which a low cost, EQAS program focusing on microscopic examination of clinical specimens was structured and launched in 2022. Performance of this national-level EQAS program over a four-year period, established following assessment of diagnostic scope and the challenges faced during implementation as well as feedback received from participating laboratories is presented. Our analysis highlights areas of strength, identifies consistent and fluctuating performers, and the program’s overall contribution to strengthening clinical parasitology diagnostics in India.

## MATERIALS AND METHODS

### Diagnostic scope assessment survey

An online survey (using SurveyCTO, Dobility, Inc; Cambridge, MA, USA) was developed to collect data on parasitology diagnostic capabilities of the participating laboratories including information on staffing, qualifications, methods, practices and scale of operations (Supplementary file 1: Form 1: Diagnostic scope assessment survey form) and circulated by email to 506 laboratories and hospitals across the country. Only a single completed response was accepted from each laboratory.

### CMC Parasitology EQAS design and workflow

The program was developed with protocols reviewed by expert advisors from international EQAS programs - UK National External Quality Assessment Service (UKNEQAS) and Stichting Kwaliteitsbewaking Medische Laboratoriumdiagnostiek (SKML) (Netherlands Foundation for Quality Assessment in Medical laboratory Diagnostics). Participation was voluntary, at no cost and open to all diagnostic laboratories from private, government run, and not-for profit hospitals. Proficiency testing (PT) parasite material were distributed biannually, with two challenges in each cycle. Before each cycle, internal planning to select samples, plan sample preparation, develop a clinical scenario with forms and handouts for distribution, and finalize the distribution schedule was conducted. Samples that met the criteria of adequate quantity (minimum of 0.5 grams per aliquot), sufficient parasite load (minimum of 3- 5 ova/smear; 3-5 cysts/high power field on direct microscopy), and morphology (based on key differentiating features and independently confirmed by two laboratory personnel) were selected. Samples were homogenized and based on the PT material provided, they were aliquoted as formalinized samples in vials, ethanol fixed stained smears, or sealed as permanent mount slides. PT material were packed according to standard practice in compliance with the UN3373 Biological Substance B criteria [11] and shipped via Indian postal services.

### EQAS Quality Control

To assess homogeneity, 10% of PT parasite material in their final packaged form were evaluated by two independent observers, who quantified the number of parasites (Supplementary file 2: Table 1: Homogeneity assessment of PT parasite material). Homogenization was considered acceptable when the parasite counts did not exceed three standard deviations from the mean. To validate the prepared PT materials, approximately 5 coded samples were provided to staff members in the diagnostic laboratory not in the EQAS team to record their findings. Any discrepancy or lack of consensus in the coded samples, required re-examination of PT materials (Supplementary file 2: Table 2: Validation of PT parasite material). The stability of the PT materials was assessed at different temperatures by incubating them at 37°C and 4°C for 24 hours and by storing two sets of PT materials at room temperature for varying lengths of time with one set assessed after 2 weeks and the other after 4 weeks. Duplicate PT materials, marked as ‘stability check’ were shipped to two distant participating laboratories in north and north-eastern India by prior agreement (>2000 km) and were returned to our institute at the end of the PT cycle for evaluation (Supplementary file 2: Table 3: Stability assessment of PT parasite material).

Prior to shipment, participants were notified by email along with an instruction sheet, safety data sheet, and a link to the reporting portal (previously SurveyCTO and currently to the CMC Parasitology EQAS website, **cmcparasiteqas.in**). Participants were given one month from the date of shipment dispatch to complete the exercise, after which the intended results were emailed. Individual participating laboratories were evaluated based on established scoring criteria (Supplementary file 1: Form 2: EQAS scoring criteria form). Each participant received a confidential report on their proficiency and a summary of the overall performance of all the participating laboratories. The PT report was also accompanied by an educational handout. To further provide educational resources and increase EQAS awareness, in October 2023, a monthly online case discussion was initiated, where case scenarios and microscopic images of parasites were shared on social media and participants were notified via email, and the results were shared a week later.

### EQAS feedback survey

After three years of the program, an online feedback survey was conducted (using a Google-doc form) in 2024 to assess the quality and user experience covering the key aspects of the program such as frequency of PT cycles, quality of the PT materials provided, timeliness/usefulness of reports/teaching materials, and EQAS team response to queries. Responses were recorded using a combination of Likert-type scales, multiple-choice questions and free-text suggestions (Supplementary file 1: Form 3: Feedback survey form).

### Statistical Analysis

The diagnostic scope assessment survey, EQAS performance and feedback survey data were analysed in Microsoft Excel (version 2108). Along with descriptive statistics for the surveys, EQAS performance ratings for each laboratory were calculated by standardized performance rating method adapted from the UK NEQAS performance assessment protocol [12]. Individual laboratory participant performance for parasites distributed was represented by heatmap analysis (R version 4.6.1; R foundation for Statistical Computing, Vienna, Austria) and included only those who had participated in more than one EQAS cycle.

### Ethical statement

The EQA program was approved by the Institutional Ethics Committee, Christian Medical College, Vellore (approval letter dated 19-Apr-2021).

## RESULTS

### Diagnostic scope assessment survey

A total of 78 laboratories participated in the survey (**Table 1**) of the 506 contacted, with a near-equal distribution between government (n=37) and private (n=41) institutions. Most diagnostic laboratories were involved in teaching (62/78, 79%), but participation in parasitology EQAS was not high (16/78, 20%). In terms of specimen types received for parasitology diagnostics, stool samples were the most common (99%), followed by blood (60%), sputum (induced sputum and bronchoalveolar lavage) (56%), pus (54%), tissue or whole worms (51%), fluid samples (51%), aspirates (47%), and corneal scrapings (40%).

**Table 1:** Laboratory Tests for Parasite Detection in India – Diagnostic Scope Assessment Survey Results.

| Parasite type | Wet mounts<br>*n* (%) | Staining techniques<br>*n* (%) | QBC<br>*n* (%) | RDTs<br>*n* (%) | Culture<br>*n* (%) | ELISA<br>*n* (%) | PCR<br>*n* (%) | Tests not done, *n* (%) |
| --- | --- | --- | --- | --- | --- | --- | --- | --- |
| **A - Intestinal Parasites [*n*=77]** |  |  |  |  |  |  |  |  |
| Amoebae | 76 (98.7) | 28 (36.3) | NA | 4 (5.1) | 4 (5.1) | 9 (11.6) | 1 (1.2) | 1 (1.2) |
| Flagellates & ciliates | 75 (97.4) | 23 (29.8) | NA | 2 (2.5) | 0 | 0 | 2 (2.5) | 1 (1.2) |
| Coccidian parasites | NA | 73 (94.8) | NA | 5 (6.4) | 0 | 1 (1.2) | 4 (5.1) | 3 (3.8) |
| Nematodes | 76 (98.7) | 0 | NA | 0 | 6 (7.7) | 0 | 0 | 0 |
| Cestodes | 74 (96.1) | 7 (9) | NA | NA | 0 | 1 (1.2) | 0 | 3 (3.8) |
| Trematodes | 63 (81.8) | 0 | NA | 0 | 0 | 0 | 0 | 15 (19.4) |
| **B - Blood Parasites** |  |  |  |  |  |  |  |  |
| Malaria [*n*= 47] | NA | 44 (93.6) | 12 (25.5) | 37 (78.7) | 0 | 0 | 2 (4.2) | 1 (2.1) |
| Lymphatic filariasis [*n*=47] | NA | 42 (89.3) | 1 (2.1) | 7 (14.8) | 0 | 1 (2.1) | 0 | 5 (10.6) |
| Leishmania [*n*=78] | NA | 54 (69.2) | 1 (1.2) | 9 (11.5) | 1 (1.2) | 0 | 1 (1.2) | 23 (29.4) |
| **C- Others [*n*=78]** |  |  |  |  |  |  |  |  |
| Tissue cestodes | 61 (78.2) | 0 | NA | 0 | 0 | 20 (25.6) * | 0 | 15 (19.2) |
| FLA | 66 (84.6) | 19 (24.3) | NA | 0 | 6 (7.6) | 0 | 1 (1.2) | 11 (14.1) |
| *Toxoplasma gondii* | 0 | 2 (2.5) | 0 | 1 (1.2) | 0 | 40 (51.2) ** | 0 | 46 (58.9) |
| *Trichomonas vaginalis* | 72 (92.3) | 20 (25.6) | NA | 0 | 2 (2.5) | 0 | 0 | 5 (6.4) |

Abbreviations: QBC: Quantitative Buffy Coat; RDT: Rapid Diagnostic Tests; ELISA: Enzyme Linked Immuno-Sorbent Assay; PCR: Polymerase Chain Reaction \*Echinococcus IgG ELISA is performed by 11 (11/78, 14.1%) laboratories and ELISA for cysticercosis antibody is performed by 9 (9/78, 11.5%) laboratories \*\* Toxoplasma IgM ELISA is performed by 21 (21/78, 26.9%), Toxoplasma IgG ELISA is performed by 16 (16/78, 20.5%) and Toxoplasma IgG avidity ELISA is performed by 3 (3/78, 3.8%) laboratories

For stool samples, 51% employed concentration methods and saline iodine wet mounts were the primary diagnostic method (76/77, 98.7%). For intestinal coccidian parasites, modified acid-fast staining was widely performed (73/77, 94.8%). Helminth diagnosis primarily involved wet mount examination, with a small proportion performing fecal egg counts, culture for *Strongyloides stercoralis* (6/77, 8% each), and gravid proglottid staining (7/77, 9%). ELISA, culture, rapid diagnostic tests (RDTs), and PCR were not used often for enteric parasites. For malaria diagnosis, microscopic examination of blood films was most common (44/47, 93.6%) followed by RDTs (37/47, 78.7%), and a small number performed quantitative buffy coat (QBC) examination (12/47, 25.5%). In diagnosing lymphatic filariasis, 89.3% (42/47) performed blood film examinations, while 15% (7/47) used RDTs. For *Leishmania* spp., the most commonly received samples were blood (83%) and bone marrow (60%) with staining and microscopy performed by 69% (54/78), followed by RDT for antibody (9/78, 11%). Hydatid IgG ELISA (11/78, 14%), and ELISA for cysticercosis antibodies (9/78, 11.5%) were performed by very few laboratories. For detection of free-living amoebae, wet mount examination was the primary diagnostic method (66/78, 85%) and culture using non-nutrient agar was carried out only in 7.6% (6/78) laboratories. *Toxoplasma gondii* ELISA was employed by 51% of laboratories, with IgM, IgG, and IgG avidity ELISA performed (in 21/78, 26.9%, 16/78, 20.5% and 3/78, 3.8% respectively). For *Trichomonas vaginalis*, saline iodine wet mounts were used by 92% (72/78) of laboratories and only 2.5% (2/78) carried out culture.

### Parasitology EQAS characteristics and performance

The program was initiated in 2022 with enrolment of 52 laboratories in the first cycle, expanding to 131 participants by the end of 2025. **Figure 1** shows the state wise distribution of participating laboratories across India. The average response rate across all the cycles was 92% (range from 87% to 96%). In terms of feasibility, breakage/leakage was reported only in 0.25 % (4/1554) of samples across all rounds. In terms of EQAS internal quality control (Supplementary file 2: Table 1: Homogeneity assessment of PT parasite material), all PT items (184 samples) evaluated for homogeneity showed consistent parasite counts within two standard deviations of the mean. No discrepancies or lack of consensus were observed during validation of 80 coded samples (Supplementary file 2: Table 2: Validation of PT parasite material). Stability checks revealed no deterioration in parasite morphology or staining quality across different temperatures, transportation/storage conditions and time points (a total of 96 samples with 32 samples in each group) (Supplementary file 2: Table 3: Stability assessment of PT parasite material).

**Figure 1:**
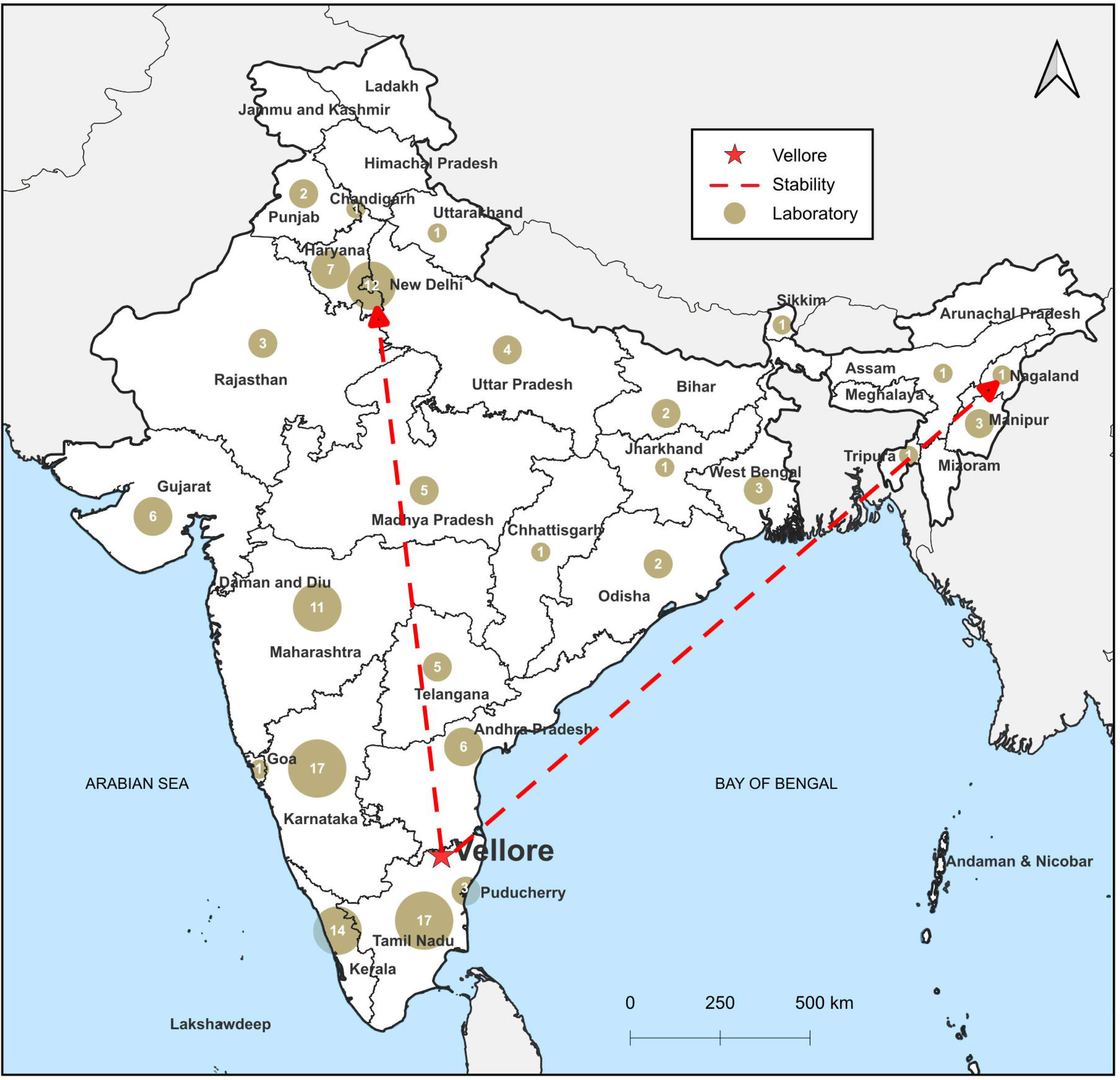
State-wise distribution of participating laboratories over four years of CMC Parasitology EQAS (2022-2025) across India (n=131) Footnote: Base map source: SimpleMaps; annotations and visualization customized in R (version 4.6.1) using ggplot2 and ggh4x packages

Overall performance of participating laboratories for individual PT challenges was variable with correct response rates ranging from 27% (26/95) to 97% (100/103) across the parasites distributed. The heat map analysis of individual laboratory performances **(Fig 2)** across eight EQAS cycles revealed that 41 laboratories achieved overall scores exceeding 90% consistently (high performers), 35 scored between 80 - 89%, 17 scored between 65 - 79%, and 20 scored constantly below 65% (poor performance). Most laboratories demonstrated high diagnostic competence in identifying routinely observed parasites accurately, as evidenced by 90% (62/69) of laboratories correctly identifying *Giardia duodenalis* cysts in cycle 2 in 2022 increasing to 97% (100/103) in cycle 7 in 2025 and 89.2% (74/83) successfully identifying both *G. duodenalis* trophozoites and cysts in stained smears in cycle 3 in 2023. Single-species helminth infections, including *Trichuris trichiura* (109/115, 95%), *Taenia spp.* ova (78/88, 88.6%), *Hymenolepis nana* (58/69, 84%) and *Strongyloides* larvae (83/95, 87%) were accurately identified by most participants. High level of competence was observed in identifying *Echinococcus granulosus* in cyst fluid mounts with 96.4% (80/83) providing correct responses. A decrease in performance scores was observed during two instances: 1) in PT cycle 5 in 2024 when a sample simulating multi-parasitic infection was distributed (27%, 26/95) and 2) PT cycle 4 in 2023 cycle when *Paracapillaria philippinensis* ova were provided (51/88, 58%). *Clonorchis sinensis* ova (75/104, 72%) during PT cycle 6 in 2024 and *Hymenolepis diminuta* ova (65/103, 63%) in PT cycle 7 in 2025 identification were also lower than 80%. In a small minority of cases, failure to specify the correct developmental stage for cysts (05/332, 1.5%) and ova (4/364, 1%) and misidentification of plant fibre/hair fragments as *Enterobius vermicularis* and *S. stercoralis* larvae (7/104, 7%) and yeast cells as *Cryptosporidium* spp. oocysts (01/50, 2%) was noted. Morphologically indistinguishable hookworm ova were also incorrectly reported with 56% (28/50) of laboratories reported *Ancylostoma* or *Necator*.

**Figure 2:**
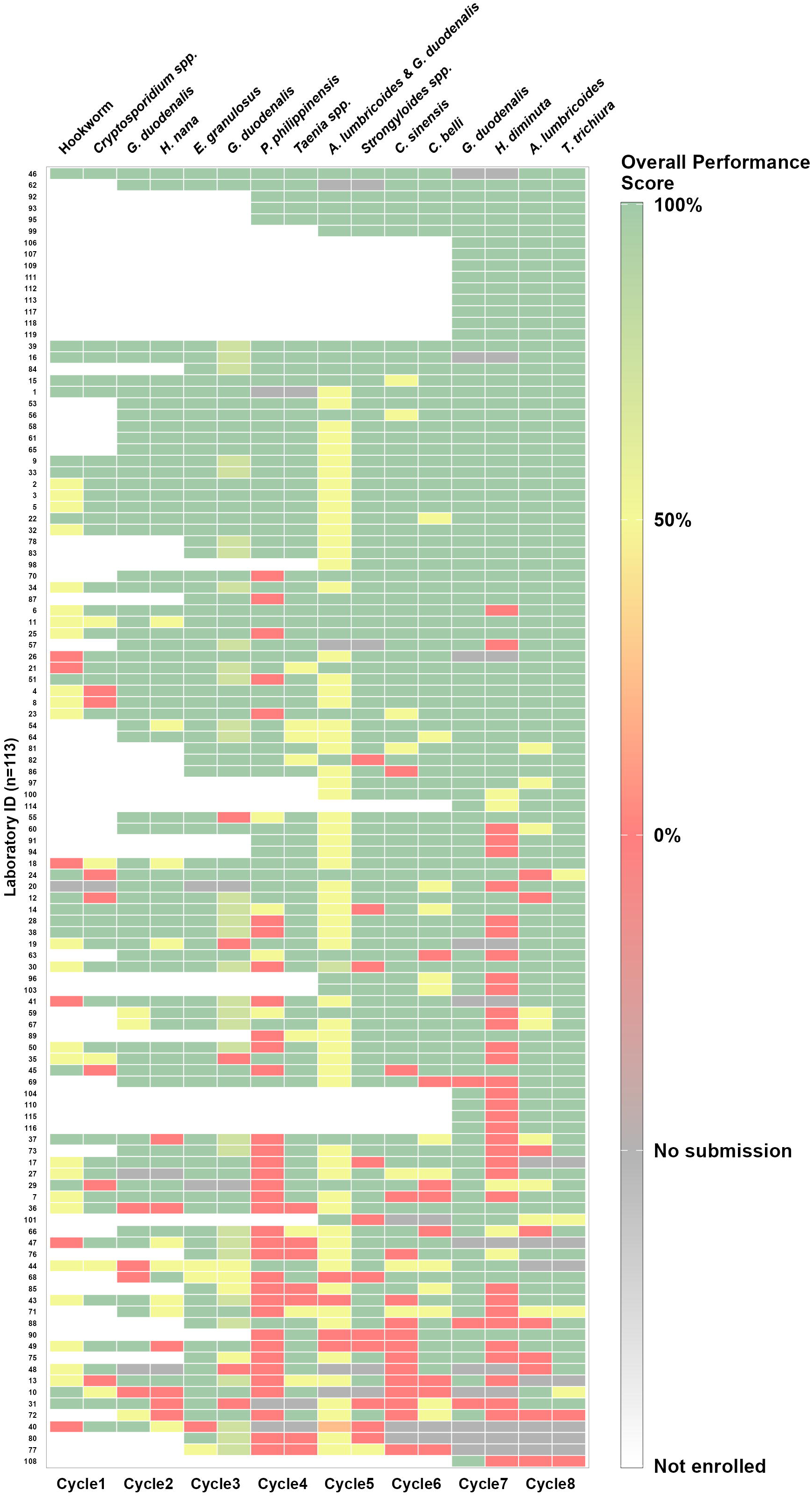
Heat map of individual participating laboratory performance for sixteen parasite challenges across eight PT cycles (2022-2025) Footnote: Each row corresponds to a participating laboratory, and each column represents a PT parasite material distributed. (For parasite material distributed see S2 Table 1)

### EQAS feedback

Three years after launching the PT program, 65 laboratories responded to the feedback survey in 2024. Nearly all respondents (96.9%) found the PT reports sufficiently informative. Although a majority of the participants (67.7%) were satisfied with the current cycle frequency, 90.8% (56/65) recommended increasing to three cycles per year in future. The quality of PT items was rated highly, with 73.8% of respondents describing them as ‘excellent’ and 24.6% as ‘good’. The challenges reported by participants included scanty distribution of *P. philippinensis* ova leading to missed identification (2/65, 3%), difficulty in identifying multiple parasites, limited experience with uncommon parasites due to infrequent encounters, and difficulty in identifying parasites correctly, each reported by one participant (1/65, 1.5%). In terms of educational resources, 98.5% (64/65) regarded the accompanying educational handout with each PT cycle as highly valuable and 63% (41/65) mentioned regular participation in the monthly online case discussions. Participants’ recommendations also included requests for periodic workshops and webinars to strengthen their diagnostic skills.

## DISCUSSION

This report summarizes the establishment and performance of a national level structured parasitology EQAS program informed by a prior assessment of diagnostic scope assessment along with feedback on improvement and future direction from participating laboratories. To the best of our knowledge, this is the first report to comprehensively assess parasitology diagnostic capabilities in Indian laboratories. Microscopy remains the most commonly employed diagnostic method across participating laboratories, underscoring that it continues to serve as the most accessible diagnostic method, especially in resource-limited settings where molecular assays and other advanced techniques are confined to research facilities. Similar findings have been reported from other studies from China, sub-Saharan Africa, Europe, and Indonesia [6, 13–15]. Although RDTs are increasingly used in malaria endemic regions, our findings indicate that microscopy remains the mainstay in diagnostic laboratories providing clinically useful data on parasitaemia, and response to treatment [16]. Prior participation in parasitology EQAS programs among the laboratories surveyed was low, highlighting a critical gap in EQAS access and awareness. This observation is consistent with findings from other low- and middle-income countries, where participation is hindered by factors such as high program costs, lack of qualified staff and the needs for a tailored EQAS with repertoire to fulfill local needs, for example, the EQAS for India would need fewer specimens of *Schistosoma* eggs and rare non-endemic parasites could be sent less frequently [17].

High accuracy was observed for common endemic parasites such as *G. duodenalis, E. granulosus, Cryptosporidium spp., S. stercoralis, H. nana, Taenia spp., A. lumbricoides* and *T. trichiura.* This aligns with earlier reports from established UK NEQAS parasitology program, where diagnostic proficiency was higher for familiar parasites [14]. In contrast, a recent European study reported misidentifications of *S. stercoralis* and *Cryptosporidium spp*., which could be attributed to shift in the diagnostic practices where traditional microscopy is replaced (e.g., dipsticks or PCR for the detection) [18]. Among these repeat measures over time, improvement in the correct identification of repeatedly distributed *G. duodenalis* was observed.

Less common parasites and parasites not endemic to India were more challenging with participants having difficulty recognizing *P. philippinensis, H. diminuta* and *C. sinensis,* as the former two are rarely encountered in routine diagnostics across most laboratories in India [19,20] and *C. sinensis* is non-endemic to most parts of the country except for rare reports from the north-eastern states [21]. Diagnostic accuracy was also lower in the sample containing two parasites, with many participants reporting only one organism. This is likely due to the tendency to discontinue further examination once a single parasite species is identified. Difficulties in reporting multiple organisms have been documented [14, 22–23]. Misidentification persisted for the duration of EQAS including misinterpreting artefacts as ova or cysts, failure to specify the correct developmental stage, and confusion between morphologically similar structures, all of which underscore the importance of ongoing competency assessment and EQAS programs.

The program faced a few logistical hurdles that impeded smooth implementation. Common issues included laboratories failing to document the correct unique identifier on return forms and outdated contact information or address changes that were not communicated, leading to delays or non-receipt of PT items and some non-responders.

Positive feedback from participants highlighted the program’s acceptability and shows that the PT initiative met the technical and learning needs for India. In response to participant suggestion, the frequency of the PT cycles will be increased to three in future and annual training activities such as workshops or webinars planned. In addition, based on our analysis, future PT will include more mixed infections and less common parasites to reflect the need to improvement in meeting real-world challenges in diagnostics. Going forward, increasing engagement with underperforming laboratories, will be a key focus. Pursuing National Accreditation Board for Testing and Calibration Laboratories (NABL) accreditation is also planned and will strengthen the PT program credibility and encourage more participation.

In conclusion, microscopy skills remain essential for accurate diagnostics in clinical parasitology especially in India, and requires continuous practice, training, and assessment. Both the Indian Council of Medical Research National Essential Diagnostic List 2025 and the Indian Public Health Standards (IPHS) Guidelines for Health and Wellness Centre–Primary Health Centre (HWC-PHC) [24,25] include microscopic examination for diagnosis of malaria, lymphatic filariasis and stool parasitic infections in primary health care settings, reinforcing the need to strengthen microscopy skills nationally. Affordable and accessible EQAS programs will help assess performance, identify areas for improvement, and ensure quality assurance in clinical parasitology.

## Supporting information

Supplemental Forms 1-3

## Data Availability

All data produced in the present work are contained in the manuscript

## Acknowledgments

We gratefully acknowledge Dr Eun Jeong Won, Asan Medical Centre, Seoul, South Korea, for providing faecal suspensions containing *Clonorchis sinensis* ova. We express our gratitude to Ms. Kavitha, and Mr. Selvakumar Prasad for statistical and data visualisation support; Ms. Elamathi and Ms. Keerthana for their contribution to sample preparation; and Ms Noel JM Hillari for enrolling new participants and maintaining the participant directory. We acknowledge the use of Editage (www.editage.com) for language, grammar improvement during manuscript preparation.

## Financial Support

This study and EQAS is funded by internal funds at The Wellcome Trust Research Laboratory, Christian Medical College, Vellore.

## Disclosures

None to declare

## Supplementary Files

Supplementary File 1: Form 1: Diagnostic Scope Assessment Survey, Form 2: EQAS Scoring Criteria, Form 3: Feedback Survey

Supplementary File 2: Table 1: Homogeneity Assessment of PT parasite material, Table 2: Validation of PT parasite material, Table 3: Stability assessment of PT parasite material

## Notes

### Competing Interest Statement

The authors have declared no competing interest.

### Author Declarations

Ethics committee/IRB of Christian Medical College Vellore gave ethical approval for this work

