## Supplemental Forms 1-3 for "Establishment of an External Quality Assessment Scheme for Clinical Parasitology Diagnostics in India: Four years of experience (2022-2025)"

**Supplementary File 1**

**S1 Form 1: Diagnostic Scope Assessment Survey Form**

**Note: The information gathered through the survey will be anonymized and used to inform the parasitology scientific community regarding the extent/measure of parasitology diagnostics in India. No laboratory/personal identifiers will be disclosed under any circumstances.**

1. Name of the institution

___________________________________

2. Name of the laboratory (or) department:

___________________________________

3. Name of the head of the laboratory (or) department:

___________________________________

4. Designation of the laboratory (or) department head

- Professor/Associate Professor/Assistant Professor
- Laboratory Supervisor
- Laboratory Technician
- Others (please specify) ____________

5. Qualification of laboratory (or) department head

- MSc
- MBBS
- MD
- PhD
- Others (please specify) ____________

6. Institution address for shipment

___________________________________

7. Name of the contact person to whom shipment should be addressed

___________________________________

8. Contact person’s phone number

Land line: __________________________

Mobile: ___________________________

9. Contact person’s e-mail

Primary: ___________________________

Alternate: ___________________________

10. Number of parasitology tests performed on average (weekly)

__________________________________

11. Years of experience in parasitology diagnostics

___________________________________

12. Laboratory’s involvement in teaching medical parasitology courses (Tick as many as applicable)

- MD
- MBBS
- MSc/BSc
- Diploma
- If any other courses (please specify) ________________
- Not applicable

13. Current participation in EQAS (Parasitology/Microbiology)

- United Kingdom National External Quality Assessment Service (UK NEQAS)
- College of American Pathologists (CAP)
- Indian Academy of Tropical Parasitology (IATP)
- Indian Association of Medical Microbiologists (IAMM)
- Others (please mention the name of the EQAS program)

________________________________

O	Not participating in EQAS

14. Types of samples received for parasitology diagnostics (Tick as many as applicable)

- Stool
- Blood
- Abscess/Pus
- Sputum/Induced sputum/Bronchoalveolar lavage (BAL)
- Fluids; O Yes O No

If yes, specify type of fluids received

O Cerebrospinal fluid

O Cyst fluid

O Pleural fluid

O Peritoneal fluid

O Urine

- Aspirates; O Yes O No

If yes, specify type of aspirates received

- Bone marrow/splenic aspirate

O Lymph node aspirate

O Bronchial aspirate

O Duodenal/Jejunal aspirate

O Bile aspirate

- Corneal scraping
- Tissue/Whole worms
- Others (please specify) ____________

15. Concentration techniques prior to fecal wet mount preparation: O Yes O No

If yes, mention the technique followed ______________________________

16. Tests for intestinal amoebae performed: O Yes O No

- Saline Iodine wet mounts: O Yes O No
- Staining techniques performed: O Yes O No

If yes, click on the stains used

O Iron hematoxylin

O Standard trichrome

O Immunofluorescence staining

O Others (please specify): ____________

- Culture: O Yes O No

If yes, mention the medium used for culture: ______________________

- Rapid diagnostic test for *Entamoeba histolytica*: O Yes O No

If yes, mention the kit used for rapid diagnostic test _________________

- Antigen detection ELISA for *Entamoeba histolytica*: O Yes O No

If yes, mention - Sample type: ___________; Kit used: ________________

- PCR for *Entamoeba histolytica*: O Yes O No; If yes, mention

Sample type: O Stool O Pus O Others (please specify) ________

Primer target: _________

- Other tests (please specify): __________________

17. Tests for intestinal flagellates & ciliates performed: O Yes O No

- Saline Iodine wet mounts: O Yes O No
- Staining techniques performed: O Yes O No

If yes, click on the stains used (Tick as many as applicable)

O Rapid Field’s stain

O Giemsa stain

O Standard trichrome stain

O Iron hematoxylin

O Immunofluorescence staining

O Other stains (please specify) _______________

- Culture for *Giardia spp*. O Yes O No

If yes, mention the medium used: _________________

- Rapid diagnostic tests for *Giardia spp.*: O Yes O No

If yes, mention the kit used: ___________________

- Antigen detection ELISA for *Giardia spp.*: O Yes O No

If yes, mention the kit used: ___________________

- PCR for *Giardia spp*.: O Yes O No

If yes, mention the sample type: O Stool O Others (please specify) __________

Primer target: _____________

- Other tests (please specify): ___________________

18. Tests for intestinal coccidian parasites

(*Cryptosporidium spp.*, *Cystoisospora belli*, *Cyclospora cayetanensis*) and Microsporidium

- Staining techniques performed: O Yes O No

If yes, click on the stains used (Tick as many as applicable)

O Modified acid-fast stain

O Calcofluor white stain

O Modified trichrome stain for Microsporidia

O Immunofluorescence staining for *Cryptosporidium spp*.

O UV fluorescence microscopy for *Cyclospora cayetanensis*

O Other stains (please specify)

- Rapid diagnostic tests for *Cryptosporidium spp.*: O Yes O No

If yes, mention the kit used _________________________

- PCR for *Cryptosporidium spp*. O Yes O No; If yes, mention

Sample type: O Stool O Intestinal Aspirates O Biopsy; Primer target: ___________

- PCR for *Cystoisospora belli* O Yes O No; If yes, mention

Sample type: O Stool O Intestinal Aspirates O Biopsy; Primer target: ___________

- PCR for *Cyclospora cayetanensis* O Yes O No; If yes, mention

Sample type: O Stool O Intestinal Aspirates O Biopsy; Primer target: ___________

- Other tests (please specify): _________________________

19. Tests for intestinal nematodes performed: O Yes O No

- Saline Iodine wet mounts: O Yes O No
- Fecal egg counts (FEC): O Yes O No

If yes, mention the method used: _________________

- Culture for *Strongyloides stercoralis*;

If yes, mention the medium used: __________________

- PCR for *Ancylostoma duodenale*	O Yes O No

If yes, mention the primer target ___________

- PCR for *Necator americanus* O Yes O No

If yes, mention the primer target _______________

- PCR for *Ascaris lumbricoides* O Yes O No

If yes, mention the primer target _______________

- PCR for *Trichuris trichiura* O Yes O No

If yes, mention the primer target _________________

- PCR for *Strongyloides stercoralis* O Yes O No

If yes, mention the primer target ___________

- PCR for *Enterobius vermicularis* O Yes O No

If yes, mention the primer target ___________

- Other tests (please specify): ______________

20. Tests for intestinal cestodes performed: O Yes O No

- Saline Iodine wet mounts: O Yes O No
- Stained gravid proglottids: O Yes O No

If yes, click on the stains used

O India ink

O Carmine

O Other stains (please specify) _____________

- PCR for *Taenia saginata* O Yes O No

If yes, mention the primer target ______________

- PCR for *Hymenolepis nana* O Yes O No

If yes, mention the primer target ______________

- PCR for *Hymenolepis diminuta* O Yes O No

If yes, mention the primer target ______________

- PCR for *Dibothriocephalus latus* O Yes O No

If yes, mention the primer target ______________

- Other tests (please specify) __________________

21. Tests for Malaria performed: O Yes O No

- Stained thick/thin blood films for microscopic examination: O Yes O No

If yes, click on the stains used

O Giemsa

O Field’s stain

O Leishman

O Other stains (please specify) __________________________

- Quantitative Buffy Coat (QBC): O Yes O No
- Rapid diagnostic tests: O Yes O No

If yes, mention the targets detected; Kits used: _______________

O Histidine rich protein-2 (HRP-2)

O pLDH

O Plasmodium aldolase

O Combined targets

- PCR for *Plasmodium vivax* O Yes O No

If yes, mention the primer target ____________

- PCR for *Plasmodium falciparum* O Yes O No

If yes, mention the primer target ____________

- PCR for *Plasmodium ovale* O Yes O No

If yes, mention the primer target ____________

- PCR for *Plasmodium malariae* O Yes O No

If yes, mention the primer target ____________

22. Tests for Leishmania performed: O Yes O No

- Stained smear for microscopic examination: O Yes O No

If yes, mention the sample type

O Blood

O Bone marrow

O Skin/tissue

O Buffy coat

Mention the stains used

O Giemsa

O Field’s stain

O Leishman

O Other stains (please specify) ___________________

- Culture methods performed O Yes O No

O Novy-MacNeal-Nicolle (NNN) culture

O Other culture methods (please specify) _______________

- Rapid diagnostic tests: O Yes O No

If yes, mention the target detected; Kits used: _______________

O rK39 antigen

O rKE16 antigen

- PCR for Leishmania: O Yes O No; If yes, mention

Sample type: O Blood O Bone Marrow O Skin/tissue O Others (please specify) __________

Primer target ___________

- Other tests (please specify) _____________________

23. Tests for lymphatic filariasis performed: O Yes O No

- Stained thick/thin blood films for microscopic examination: O Yes O No

If yes, click on the stains used

O Giemsa

O Field’s stain

O Leishman

O Other stains (please specify) ___________________

- Quantitative Buffy Coat (QBC): O Yes O No
- Filariasis test strip for circulating filarial antigen: O Yes O No

If yes, specify the kit used: ______________________

- PCR: O Yes O No; If yes, mention

Sample type: O Blood O Dried blood spots O Others (please specify) ___________

Primer target___________

- Other tests (please specify) _________________

24. Tests for tissue cestodes performed: O Yes O No

- Hydatid fluid wet mounts: O Yes O No
- Echinococcus IgG ELISA: O Yes O No

If yes, mention the kit used: _______________________

- Hydatid PCR: O Yes O No

If yes, mention the primer target ____________________

- Other tests (please specify) ________________________

25. Tests for free living amoeba performed: O Yes O No

- Wet mount microscopy O Yes O No
- Stained smears O Yes O No

If yes, click on the stains used (Tick as many as applicable)

O Lactophenol cotton blue

O Calcofluor white

O Giemsa

O Other stains (please specify) __________________________

- Culture: O Yes O No; If yes, mention the method used for culture ______________
- PCR for *Acanthamoeba spp*. O Yes O No; If yes, mention

Sample type: O CSF O Corneal scraping O Others (please specify) __________

Primer target: ________________

- PCR for *Naegleria fowleri O* Yes O No; If yes, mention

Sample type: O CSF O Brain tissue O Others (please specify) ___________

Primer target: ________________

- PCR for *Balamuthia mandrillaris* O Yes O No; If yes, mention

Sample type: O Tissue O Others (please specify) __________; Primer target: _____________

- Other tests (please specify) _______________________

26. Tests for trematodes performed: O Yes O No

- Saline Iodine wet mounts: O Yes O No
- PCR for *Fasciola spp.* O Yes O No; If yes, mention

Sample type: O Stool O Bile O Aspirates

Primer target: ___________

- PCR for *Paragonimus westermanii* O Yes O No; If yes, mention

Sample type: O Sputum O Stool O Aspirates

Primer target: _________

- PCR for *Schistosoma spp*. O Yes O No; If yes, mention

Sample type: O Stool O Urine O Bile O Aspirates

Primer target____________

- Other tests (please specify) _____________________

27. Tests for *Toxoplasma gondii* performed*:* O Yes O No

- Toxoplasma IgM ELISA: O Yes O No

If yes, mention the kit used: _______________

- Toxoplasma IgG ELISA: O Yes O No

If yes, mention the kit used: _______________

- Toxoplasma IgG avidity ELISA: O Yes O No

If yes, mention the kit used: ________________

- PCR for *Toxoplasma gondii:* O Yes O No

If yes, mention the sample type: O CSF O Blood O Tissue samples

Primer target: __________

- Other tests (please specify) ________________

28. Tests for *Trichomonas vaginalis* performed: O Yes O No

- Wet mounts O Yes O No
- Staining techniques performed: O Yes O No

If yes, click on the stains used

O Giemsa

O Modified Field’s stain

O Acridine orange

O Papanicolaou stain

O Immunofluorescence staining

O Other stains (please specify) _______________

- InPouch TV culture O Yes O No
- Other culture methods: O Yes O No

If yes, mention the medium used: _____________

- Other tests (please specify): __________________

Submit

**S1: Form 2: EQAS Scoring Criteria Form**

| **Criteria** | **Marks awarded** |
| --- | --- |
| Correct identification (name and stage of the parasite) | 2 |
| Inadequate identification (incorrect species or stage reported) | 1 |
| Unexpected parasite reported in addition to intended parasite | 1 |
| Incorrect identification (or) intended parasite not reported | 0 |
| Negative specimens (those not containing parasites) | |
| Correct result of no parasite found | 2 |
| Unexpected parasite reported | 0 |

Two marks awarded for each parasite; A maximum of three parasite species can be included

per proficiency testing item

**S1: Form 3: Feedback Survey form**

- Laboratory ID:
- Institute/hospital name:
- Do you find the frequency of the PT cycles sufficient?

☐ Appropriate frequency ☐ Too frequent ☐ Not frequent enough

- If you think the frequency should change from the current 2cycles/year, what do you suggest?

☐ 1 cycle/year	☐ 3 cycles/year	☐ 4 cycles/year

- How would you rate the quality of the **PT items** provided?

☐ Excellent ☐ Good ☐ Average ☐ Poor

If average/poor, why

- How clear and easy to follow were the **instructions** provided for submitting results?

☐ Easy ☐ Neutral ☐ Can be improved ☐ Difficult ☐ Confusing

If selected “Can be improved/Difficult/Confusing”, please provide your suggestions

- Is the information provided in the **PT report** sufficient to assess your laboratory’s performance?

☐ Sufficient ☐ Can be improved ☐ Insufficient

If selected “Can be improved/ Insufficient”, please provide your suggestions

- Were the reports/results provided in a timely manner?

☐ Yes ☐ No

- How useful do you find the **teaching materials** provided?

☐ Very useful ☐ Somewhat useful ☐ Not useful

If selected “Somewhat useful/Not useful”, please provide your suggestions

- Were your **queries or concerns** adequately addressed by the EQAS team?

☐ Addressed on time ☐ Can be improved ☐ Not addressed

If selected “Can be improved”, please provide your suggestions

- What challenges (if any) did you face during the program?
  ☐ Sample handling ☐ Submission of results ☐ Communication with the team
  ☐ Others:
- Are you participating in the **monthly twitter challenge**?

☐ Yes ☐ No

- Would you still participate if the program required a nominal fee for sustainability?

☐ Yes ☐ No ☐ Unsure

- How would you rate the **overall experience** with the program?
   ☐ Excellent ☐ Very Good ☐ Good ☐ Average ☐ Poor
- Additional comments or suggestions for improvement:

___________________________________________________________________

Thank you for your valuable feedback!
