## Supplemental Tables 1-3 for "Establishment of an External Quality Assessment Scheme for Clinical Parasitology Diagnostics in India: Four years of experience (2022-2025)"

**Supplementary File 2**

**S2: Table 1: Homogeneity Assessment of Proficiency Testing (PT) Parasite Material**

| **PT cycle** | **PT material** | **Parasite/field** | **PT material format provided*** | **Observer 1 (Mean)** | **Observer 2 (Mean)** | **Overall Mean** | **SD** | **Homogeneity status**^#^ |
| --- | --- | --- | --- | --- | --- | --- | --- | --- |
| 1 | 2022/A1 | Hookworm ova/smear | FFS in vial | 40.4 | 41 | 40.7 | 3.2 | Homogenous |
|  | 2022/A2 | *Cryptosporidium spp.,* oocysts/OIF | Modified acid-fast stained slide | 80.7 | 82.5 | 81.6 | 5.95 | Homogenous |
| 2 | 2022/B1 | *Giardia duodenalis* cyst/HPF | FFS in vial | 8.6 | 9 | 8.8 | 1.32 | Homogenous |
|  | 2022/B2 | *Hymenolepis nana* ova/smear | FFS in vial | 21.6 | 20.7 | 21.1 | 1.84 | Homogenous |
| 3 | 2023/A1 | *Echinococcus granulosus* hooklets & protoscolices/HPF | Cyst fluid in permanent mount | 3.9 | 3.8 | 3.85 | 0.92 | Homogenous |
|  | 2023/A2 | *Giardia duodenalis* trophozoites & cysts/HPF | Trichrome stained slide | 1.3 & 3 | 1.4 & 3 | 1.35 & 3 | 0.49 & 0 | Homogenous |
| 4 | 2023/B1 | *Paracapillaria philippinensis* ova/smear | FFS in vial | 6.9 | 7.3 | 7.1 | 0.79 | Homogenous |
|  | 2023/B2 | *Taenia spp.,* ova/smear | FFS in vial | 11.3 | 10.6 | 10.9 | 1.40 | Homogenous |
| 5 | 2024/A1 | *Ascaris lumbricoides* ova/smear | FFS in vial | 7.2 | 7.5 | 7.35 | 0.67 | Homogenous |
|  |  | *Giardia duodenalis* cyst/HPF |  | 4.2 | 4.4 | 4.3 | 0.98 |  |
|  | 2024/A2 | *Strongyloides* larvae/HPF | FFS in vial | 30.6 | 30.3 | 30.45 | 2.09 | Homogenous |
| 6 | 2024/B1 | *Clonorchis sinensis* ova/smear | Stool mount | 11.5 | 11.8 | 11.65 | 1.46 | Homogenous |
|  | 2024/B2 | *Cystoisospora belli* oocysts/OIF | Modified acid-fast stained slide | 39.9 | 41.1 | 40.5 | 2.98 | Homogenous |
| 7 | 2025/A1 | *Giardia duodenalis* cysts/HPF | FFS in vial | 7.35 | 7.8 | 7.5 | 0.94 | Homogenous |
|  | 2025/A2 | *Hymenolepis diminuta* ova/smear | FFS in vial | 11.2 | 12.7 | 11.9 | 1.47 | Homogenous |
| 8 | 2025/B1 | *Ascaris lumbricoides* ova/HPF | Stool mount | 9.9 | 9.2 | 9.55 | 1.29 | Homogenous |
|  | 2025/B2 | *Trichuris trichiura* ova/smear | Stool mount | 13.5 | 13.8 | 13.65 | 1.96 | Homogenous |

Abbreviations - FFS: Formalinized Fecal Suspension; HPF: High Power Field; LPF: Low Power Field; OIF: Oil Immersion Field;

SD: Standard Deviation

#PT material was considered homogenous when the parasite count was within ± 3 SD of the mean

**S2: Table 2: Validation of Proficiency Testing (PT) Parasite Material**

| **PT Material** | **Expected result*** | **Consensus result (n/5)** | **Agreement^#^** |
| --- | --- | --- | --- |
| 2022/A1 | Hookworm ova | (5/5) | 100% |
| 2022/A2 | *Cryptosporidium spp.,* oocysts | (5/5) | 100% |
| 2022/B1 | *Giardia duodenalis* cyst | (5/5) | 100% |
| 2022/B2 | *Hymenolepis nana* ova | (5/5) | 100% |
| 2023/A1 | *Echinococcus granulosus* hooklets & protoscolices | (5/5) | 100% |
| 2023/A2 | *Giardia duodenalis* trophozoites & cysts | (5/5) | 100% |
| 2023/B1 | *Paracapillaria philippinensis* ova | (5/5) | 100% |
| 2023/B2 | *Taenia spp.,* ova | (5/5) | 100% |
| 2024/A1 | *Ascaris lumbricoides ova & G. duodenalis* cyst | (5/5) | 100% |
| 2024/A2 | *Strongyloides* larvae | (5/5) | 100% |
| 2024/B1 | *Clonorchis sinensis* ova | (5/5) | 100% |
| 2024/B2 | *Cystoisospora belli* oocysts | (5/5) | 100% |
| 2025/A1 | *Giardia duodenalis* cysts | (5/5) | 100% |
| 2025/A2 | *Hymenolepis diminuta* ova | (5/5) | 100% |
| 2025/B1 | *Ascaris lumbricoides* ova | (5/5) | 100% |
| 2025/B2 | *Trichuris trichiura* ova | (5/5) | 100% |

#Agreement was defined as concordance between the expected and observed results for a given PT material. If findings were not in consensus (or) more than 20% discrepancy observed, the PT materials were re-evaluated

**S2: Table 3: Stability Assessment of Proficiency Testing (PT) Parasite Material**

| **PT material** | **PT material provided** | **37°C**  **(24 hours)** | **4°C**  **(24 hours)** | **RT**  **(2 weeks)** | **RT**  **(4 weeks)** | **Transport stability** | **Stability status**^#^ |
| --- | --- | --- | --- | --- | --- | --- | --- |
| 2022/A1 | Hookworm ova (FFS in vial) | Positive | Positive | Positive | Positive | Concordant | Stable |
| 2022/A2 | *Cryptosporidium spp.* oocysts (stained smear) | Positive | Positive | Positive | Positive | Concordant | Stable |
| 2022/B1 | *G. duodenalis* cyst (FFS in vial) | Positive | Positive | Positive | Positive | Concordant | Stable |
| 2022/B2 | *H. nana* ova in (FFS in vial) | Positive | Positive | Positive | Positive | Concordant | Stable |
| 2023/A1 | *E. granulosus* hooklets/protoscolices (formalinized cyst mount) | Positive | Positive | Positive | Positive | Concordant | Stable |
| 2023/A2 | *G. duodenalis* trophozoites and cysts (stained smear) | Positive | Positive | Positive | Positive | Concordant | Stable |
| 2023/B1 | *Capillaria philippinensis* ova (FFS in vial) | Positive | Positive | Positive | Positive | Concordant | Stable |
| 2023/B2 | *Taenia spp.,* ova (FFS in vial) | Positive | Positive | Positive | Positive | Concordant | Stable |
| 2024/A1 | *A. lumbricoides ova + G. duodenalis* cyst (FFS in vial) | Positive | Positive | Positive | Positive | Concordant | Stable |
| 2024/A2 | *Strongyloides* larvae (FFS in vial) | Positive | Positive | Positive | Positive | Concordant | Stable |
| 2024/B1 | *Clonorchis sinensis* ova (stool mount) | Positive | Positive | Positive | Positive | Concordant | Stable |
| 2024/B2 | *Cystoisospora belli* oocysts (stained smear) | Positive | Positive | Positive | Positive | Concordant | Stable |
| 2025/A1 | *G. duodenalis* cysts (FFS in vial) | Positive | Positive | Positive | Positive | Concordant | Stable |
| 2025/A2 | *H. diminuta* ova (FFS in vial) | Positive | Positive | Positive | Positive | Concordant | Stable |
| 2025/B1 | *Ascaris lumbricoides* ova (stool mount) | Positive | Positive | Positive | Positive | Concordant | Stable |
| 2025/B2 | *Trichuris trichiura* ova (stool mount) | Positive | Positive | Positive | Positive | Concordant | Stable |

Abbreviation – FFS: Formalinized Fecal Suspension

\# Stable: PT material was considered stable when the parasite identity remained unchanged, morphology of the parasite was not distorted, number of the parasites was within 3 standard deviations of the mean recorded. For stained preparations, the staining quality without fading or artefacts was considered satisfactory and for mounted slides, absence of drying and slide integrity was checked.
